# Direct detection and quantification of *Mycobacterium tuberculosis* from clinical samples by high-resolution melt qPCR

**DOI:** 10.64898/2026.03.07.26347851

**Authors:** Aishwarya S. Babu, Kalaiarasan Ellappan, Priyanka Yadav, Parthasarathy Ponnusamy, Vishnukanth Govindaraj, Saka Vinod Kumar, Akhilesh Pandey, Noyal Mariya Joseph, Renu Verma

## Abstract

**Background:** Estimating bacterial load in clinical samples has important applications in tuberculosis (TB) management, including assessment of disease severity, treatment response, and baseline genome copies required for sequencing. However, rapid and affordable tools for quantifying *Mycobacterium tuberculosis* (*M. tuberculosis*) remain limited.

**Methods:** We developed a high-resolution melt (HRM) based quantitative PCR assay using molecular beacon chemistry targeting the single-copy RD9 region of *M. tuberculosis*. Analytical performance was assessed using 10-fold serial dilutions of H37Rv DNA. Clinical validation included DNA from 100 *M. tuberculosis* culture isolates and 40 sputum samples from Xpert MTB/RIF- and culture-positive pulmonary TB patients. To evaluate specificity, we tested 30 non-tuberculous mycobacterium (NTM) culture isolates from patients infected with *Mycobacterium abscessus* (n=25) and *Mycobacterium fortuitum* (n=5) and DNA from saliva samples of 10 healthy controls.

**Results:** The HRM-qPCR assay showed a linear dynamic range from 10^1^ to 10^6^ genome copies per reaction, with a lower limit of detection of 10 copies. Standardized melt-curve analysis yielded a single target-specific peak at 73.7±0.12°C across dilutions, confirming specific amplification. Sensitivity for *M. tuberculosis* detection was 100% in culture isolates and 95.0% (38/40) in sputum, with no false positives among *M. tuberculosis*-negative controls. Assay specificity was 100% in both culture isolates and sputum, with no additional melt peaks. We did not observe any peaks indicative of non-specific binding in NTMs. Amplification was observed in two NTM samples whose Tm matched *M. tuberculosis* RD9 (median *M. tuberculosis* copies=678.5), suggesting possible co-infection or contamination. No amplification or specific melt peaks were observed in saliva samples, indicating high probe specificity.

**Conclusions:** Molecular beacon-based HRM-qPCR assay enables rapid, highly specific quantification of *M. tuberculosis* genome copies in clinical samples and has potential utility for treatment monitoring, triaging specimens for sequencing, and assessing transmission risk in high-burden settings.

## Introduction

Tuberculosis (TB) remains one of the leading causes of mortality from a single infectious agent, claiming around 1.23 million lives every year [1]. India alone accounts for ∼27% of all global TB cases, underscoring its position as the country with the greatest disease burden. *M. tuberculosis*, the etiological agent of TB, is detected in clinical samples across a wide spectrum of bacillary load, ranging from smear-negative, paucibacillary disease to smear-positive disease with very high bacillary counts. Bacterial load in a clinical sample provides crucial information on disease severity, is used to monitor treatment response, and helps assess the risk of relapse.

Culture-based quantification relies on reporting CFU/mL on solid media, which requires BSL-3 infrastructure and several weeks to obtain results, thereby limiting its scalability [2]. Additionally, culture-based methods for determining bacterial load in clinical specimens are vulnerable to contamination with other microorganisms present in the sample [3]. Furthermore, studies have shown that certain *M. tuberculosis* populations do not grow on artificial media, leading to underestimation of the true bacterial load [4]. In contrast, qPCR-based diagnostic assays for *M. tuberculosis* are highly sensitive, do not get impacted by contamination or presence of culturable or non-culturable bacteria and can be performed directly on DNA extracted from sputum or other clinical specimens. However, the qPCR assays used in routine practice are predominantly qualitative or semiquantitative and therefore do not provide absolute bacterial load measurements in patient samples.

An assay that measures absolute *M. tuberculosis* copies in clinical samples would therefore be valuable for both patient management and research. Several studies underscore this need. The TB molecular bacterial load assay (TB-MBLA), which quantifies *M. tuberculosis* RNA, has been used to monitor treatment response. Serial quantification of bacterial load using TB-MBLA at baseline, 2 weeks and 2 months distinguished responders from non-responders and prompted regimen changes or adherence interventions [5]. Additionally, molecular bacterial load assays are increasingly being adopted as endpoints in clinical trials, where absolute load and its early decline over 14-56 days serve as key markers for evaluating new regimens [6]. When combined with clinical and aerosol data, quantitative assays have been shown to improve prediction of relapse risk, refine infectiousness estimates and guide decisions on isolation and contact tracing [7].

Another important application of quantifying bacterial load in a clinical sample is prior to targeted sequencing. Targeted next-generation sequencing (tNGS) for drug-resistant TB (DR-TB), recently recommended by WHO, can detect resistance to multiple drugs in a single test, but typical per-sample costs in LMIC laboratories are around INR 15,000 (US$160), including library preparation and sequencing consumables. Establishing the minimum *M. tuberculosis* genome copy number required for successful tNGS and robust resistance calling is therefore critical. Absolute qPCR-based assays can serve as a cost-effective screening tool to identify specimens with adequate bacillary load and support efficient sequencing workflows. Beyond treatment monitoring and tNGS triage, quantitative *M. tuberculosis* load has further applications, for example, identifying patients at the highest transmission risk by correlating high bacillary load with smear positivity and infectiousness [8]. Bacterial load assessment also serves as a pharmacodynamic marker in clinical and programmatic studies to compare regimen bactericidal activity, optimize treatment duration and predict relapse [9]. It is also used to stratify baseline disease severity and prognosis and to inform decisions on hospitalisation, respiratory isolation and the intensity of follow-up [6].

Despite these applications, fully quantitative qPCR assays for *M. tuberculosis* are still largely confined to research settings, and commercially available tests remain very expensive [10,11]. For example, the MTB Complex Bacterial Load Test offered in India by DNA Labs India costs INR 5,100-6,000 (US$55-65) per sample, with a reported turnaround time of about 36 hours, including email reporting by the third working day [12]. These costs and workflow requirements make absolute-quantification assays difficult to scale in high-burden countries such as India. Additionally, most existing assays use TaqMan probe chemistry, which shows increased variability and reduced reliability near the limit of detection (LOD) because of stochastic sampling, template complexity and background signal. Furthermore, many qPCR assays for *M. tuberculosis* detection target the multicopy insertion sequence IS6110, whose copy number varies widely between strains, thereby limiting the accuracy of any inferred correlation between cycle threshold (Ct) values and true bacillary load [11,13].

In this study, we developed a molecular beacon-based qPCR assay that combines asymmetric PCR with probe-target-based HRM curve analysis for the detection of *M. tuberculosis* in clinical samples. The molecular beacon chemistry enables HRM curve analysis, generating an *M. tuberculosis*-specific melt profile that improves assay specificity in complex matrices such as sputum. We further incorporated absolute quantification by constructing standard curves. The assay was validated using sputum and culture-derived *M. tuberculosis* from patients with active TB. We also validated the specificity of our assay against NTM.

## Methods

### RD9-specific molecular beacon design

For *M. tuberculosis*-specific molecular beacon design, we targeted Region of Difference 9 (RD9), a single-copy locus in *M. tuberculosis* clinical strains that is absent from *Mycobacterium bovis* and several other *Mycobacterium tuberculosis* complex (MTBC) members and is therefore widely used as a species-specific marker. Primers and probes were manually designed to amplify a 115 bp region within RD9, and in-silico validation was performed using Beacon Designer (version 8.21). A 23-bp molecular beacon probe with 6-bp stems at both 5′ and 3′ ends was designed. Candidate probes were screened in silico to minimize secondary structure formation and non-specific interactions. Given the high GC content (∼65%) of the *M. tuberculosis* genome, regions with excessively high GC content were avoided during target selection. The 5′ end of the probe was labelled with FAM (emission maximum 517-520 nm) and the 3′ end with the dark quencher BHQ1. Finally, a universal nucleotide BLAST search was performed to exclude non-specific binding, after which the probe and corresponding primers were finalized. The following primer and probe sequences were used: RD9-forward primer 5′-AGCCTGCTGACTCATCTG-3′, RD9-reverse primer 5′-CCGATCCGTAGACATAGTTG-3′, and RD9 molecular beacon probe 5′-FAM-CGGCGCGATCGCTGGTGGTGTTCTCCTCGGCGCCG-BHQ1-3′ (Fig. 1).

**Figure 1:**
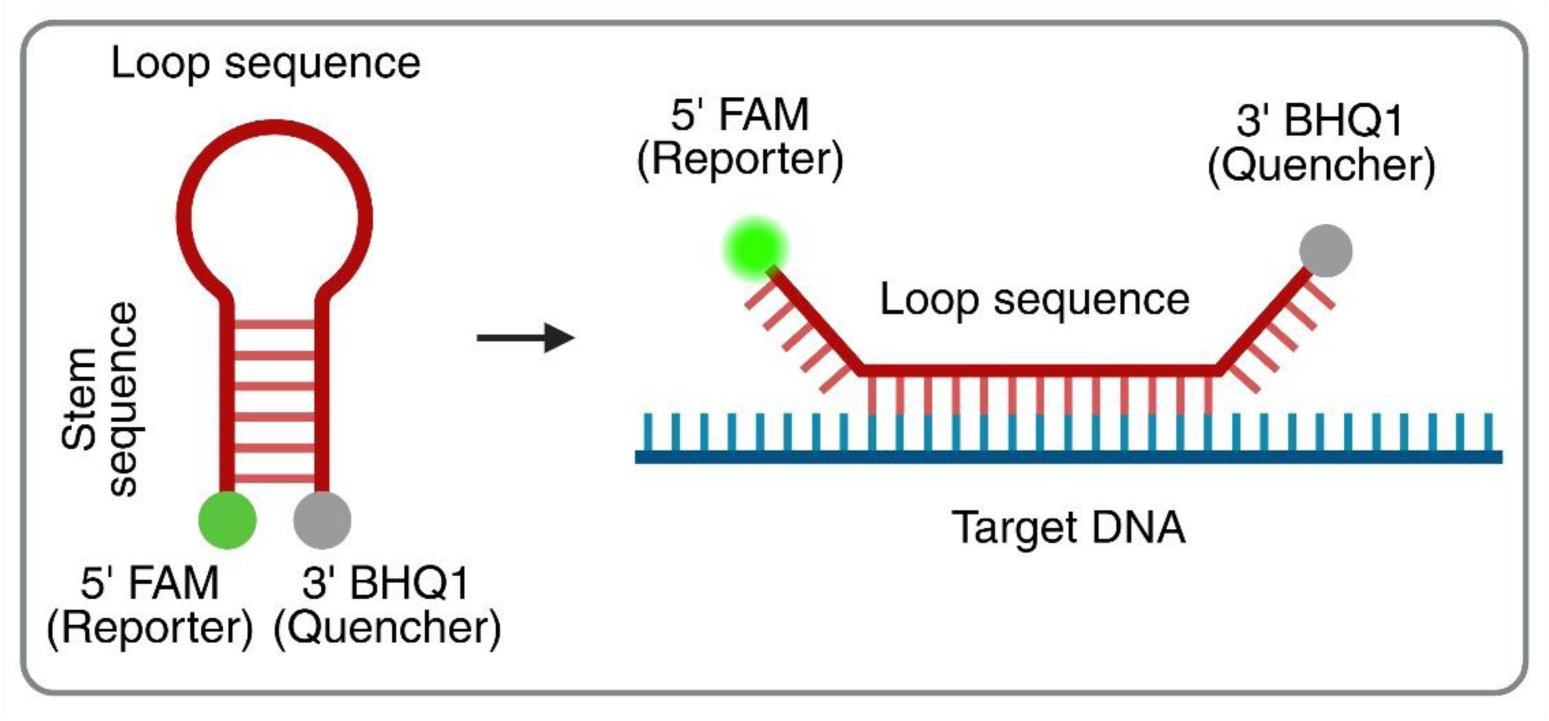
Schematic representation of the hairpin-loop structure and working principle of a molecular beacon probe. The probe forms a stem-loop structure in its native state, with a fluorescent reporter (FAM) attached at the 5′ end and a quencher (BHQ1) at the 3′ end. In the absence of a target sequence, the stem hybridises, bringing the reporter and quencher together and quenching fluorescence. Upon hybridisation of the loop sequence to its complementary target DNA, the stem structure dissociates, separating the reporter from the quencher and resulting in fluorescence emission.

### Accuracy and limit of detection assessment

Using the *M. tuberculosis* H37Rv (ATCC #25618DQ) reference strain, we performed asymmetric PCR with Vent (exo-) DNA polymerase (New England Biolabs). The qPCR mix contained 0.06 µM forward primer, 1 µM reverse primer, 0.25 µM molecular beacon probe and 1X ThermoPol Reaction Buffer, in a final reaction volume of 20 µL with ROX dye as the passive reference. The PCR was performed in technical replicates. Thermocycling conditions included an initial hold at 50°C for 2 min and 94°C for 2 min, followed by 40 cycles of denaturation at 95°C for 15sec and annealing/extension at 60°C for 1 min. Melt-curve analysis was then performed with a 2-min step at 95°C, a 1-min step at 50°C, and a gradual increase to 95°C at 0.15°C/min. Additionally, we performed limit of detection assay by preparing a ten-fold serial dilution of *M. tuberculosis* H37Rv DNA ranging from 10^6^ to 10^1^ copies to generate standard curves. All the dilutions were run in triplicate. The qPCR assays were run on a QuantStudio 5 real-time PCR system (Applied Biosystems #2725212186).

### Evaluation of RD9 assay specificity

The specificity of the RD9 assay was evaluated using human saliva samples (n=10) and NTM culture isolates (n=30). Saliva samples were collected from healthy volunteers with no clinical history of TB and DNA was extracted using the QIAamp DNA Blood Mini Kit (QIAGEN #51104) according to the manufacturer’s instructions. DNA from confirmed NTM samples was extracted using standard CTAB method. Specificity was determined based on the amplification observed in saliva samples, NTM, and NTC (no template control) reactions. HRM curve analysis was performed to confirm the presence of specific peak corresponding to the expected RD9 target-probe hybrid. In samples where late Ct values (>38) were observed, only those with well-defined melt peaks were reported as positive, whereas samples with a broad melt peak were classified as “indeterminate.”

### Ethical approval and sample collection

Prospective samples were collected after ethical approval (IOB-No. IOB/2nd IEC/2023; JIPMER-JIP/IEC-OS/332/2023). All participants were 18 years and above and provided written informed consent. We collected a total of 140 *M. tuberculosis* samples, consisting of culture isolates (n=100) and sputum samples (n=40) from Xpert MTB/RIF and culture positive pulmonary TB patients and 30 NTM culture isolates from Jawaharlal Institute of Postgraduate Medical Education & Research (JIPMER), Puducherry, between 2024-2025. The NTM sample collection was approved by the Institute Ethics Committee (Approval No. JIP/IEC-OS/265/2023-dated 03.10.2023), JIPMER, Puducherry, India.

### Sputum processing and *M. tuberculosis* DNA extraction

Approximately 2 mL of early morning sputum was collected from Xpert MTB/RIF positive patients from JIPMER. The sputum sample was divided into two equal parts. Approximately 1 mL was used for direct *M. tuberculosis* DNA extraction and the remaining 1 mL was used for culture.

*M. tuberculosis* DNA extraction from sputum was optimized in-house. Briefly, 1 mL of sputum was liquefied with 500 µL of 0.1% dithiothreitol and vortexed, then centrifuged at 10,000 rpm for 10 min. The pellet was decontaminated with equal volumes of 1 M NaOH and NaCl, washed with 1 mL PBS (pH 7.4), resuspended in 120 µL PBS and incubated at 95°C for 30 min. Chemical lysis was performed with QIAamp lysis buffers: 240 µL ALT buffer and 40 µL proteinase K (56°C, 60 min), then 400 µL AL buffer (70°C, 10 min). Mechanical disruption was added by vortexing with 200 µL of 0.1 mm zirconium beads for 2 min. After lysis, 650 µL of supernatant was transferred to a fresh tube, mixed with 325 µL of 100% ethanol, and incubated for 3 min at room temperature. The column was washed with 500 µL AW1 (8000 rpm, 1 min) and 500 µL AW2 (13,000 rpm, 3 min). DNA was eluted with 50 µL nuclease-free water after a 5-min incubation and centrifugation at 8000 rpm for 1 min and stored at -20°C until use.

### DNA extraction from *M. tuberculosis* culture

DNA was extracted from *M. tuberculosis* culture isolates using a previously optimized CTAB method [14]. Briefly, a loopful of culture from Lowenstein-Jensen (LJ) slants was suspended in 500 µL TE buffer and heat-inactivated at 95°C for 30 min, followed by snap cooling on ice. Lysozyme (20 mg/mL) and RNase (10 mg/mL) were added for cell wall lysis and degrade RNA, respectively. The mixture was incubated at 37°C for 2 hours. Subsequently, 10% SDS and proteinase K (10 mg/mL) were added for protein digestion and further lysis, followed by incubation at 65°C for 30 min. To remove carbohydrates, 5 M NaCl and CTAB-NaCl solution were added, and the mixture was incubated at 65°C for 30 min. Chloroform:isoamyl alcohol (24:1) was used for protein removal by centrifugation at 14,000 rpm for 15 min. The aqueous phase was collected, and DNA was precipitated with 0.6 volumes of ice-cold isopropanol. The pellet was washed twice with 70% ethanol, air-dried, and resuspended in 50 µL nuclease-free water. Extracted DNA was stored at -20°C until further use.

### Line probe assay Genotype Mycobacterium CM (LPA-CM) for NTM samples

A total of 30 previously stored NTM strains available in the Department of Microbiology, JIPMER, Puducherry, were included in this study. All NTM strains were revived and subcultured in MGIT 960. NTM-positive MGIT cultures that were found to be AFB positive, Ziehl-Neelsen staining, and negative for the MPT64 antigen were further subjected to the GenoType Mycobacterium CM assay (Hain Lifescience, Germany) for species identification [15]. Briefly, 1.5 mL of MGIT culture was transferred into a 2mL micro centrifuge tube and centrifuged at 10,000 rpm for 10 min. The supernatant was discarded, and DNA was extracted from the pellet using the GenoLyse kit (Hain Lifescience, Germany) according to the manufacturer’s instructions. The extracted DNA was added to the kit reagents for PCR amplification. The amplified products were subsequently subjected to reverse hybridisation on nitrocellulose membrane strips for species identification. Hybridisation and detection were carried out using the TwinCubator system (Hain Lifescience, Germany).

### Data analysis

Analytical validation included standard curve generation and determination of the LOD using serial DNA dilutions. The LOD was defined as the lowest concentration detected in ≥95% of replicates. Assay precision and reproducibility were evaluated by calculating intra- and inter-run coefficients of variation (CV%) of Ct values. Analytical specificity was assessed using NTM strains and *M. tuberculosis-*negative saliva samples. Clinical performance was evaluated by comparing qPCR results with culture-confirmed samples to determine diagnostic sensitivity. Data were analysed using R software (version 4.5.1).

### Statistical analysis

We evaluated the association between AFB smear grade and qPCR Ct values in sputum samples, coding smear as an ordinal variable (Scanty-3+) and analysing Ct as a continuous variable. After excluding missing smears, we applied Spearman’s rank correlation and summarised smear-specific Ct distributions by median values.

## Results

### Clinical cohort characteristics

For clinical validation, we used DNA from 40 sputum samples and 100 *M. tuberculosis* culture isolates collected at JIPMER Hospital. The clinical characteristics of the patients are summarised in Table 1. The median age of TB patients was 50 years (range 42-55 years), and approximately 70% were men. Smear information was available for 120 pulmonary TB samples. Among those 10% (12/120) were graded 3+, 30% (36/120) were 2+, 56.6% (68/120) were 1+, and 3.3% (4/120) were scanty. All TB samples included in this analysis were from new cases.

**Table 1:**
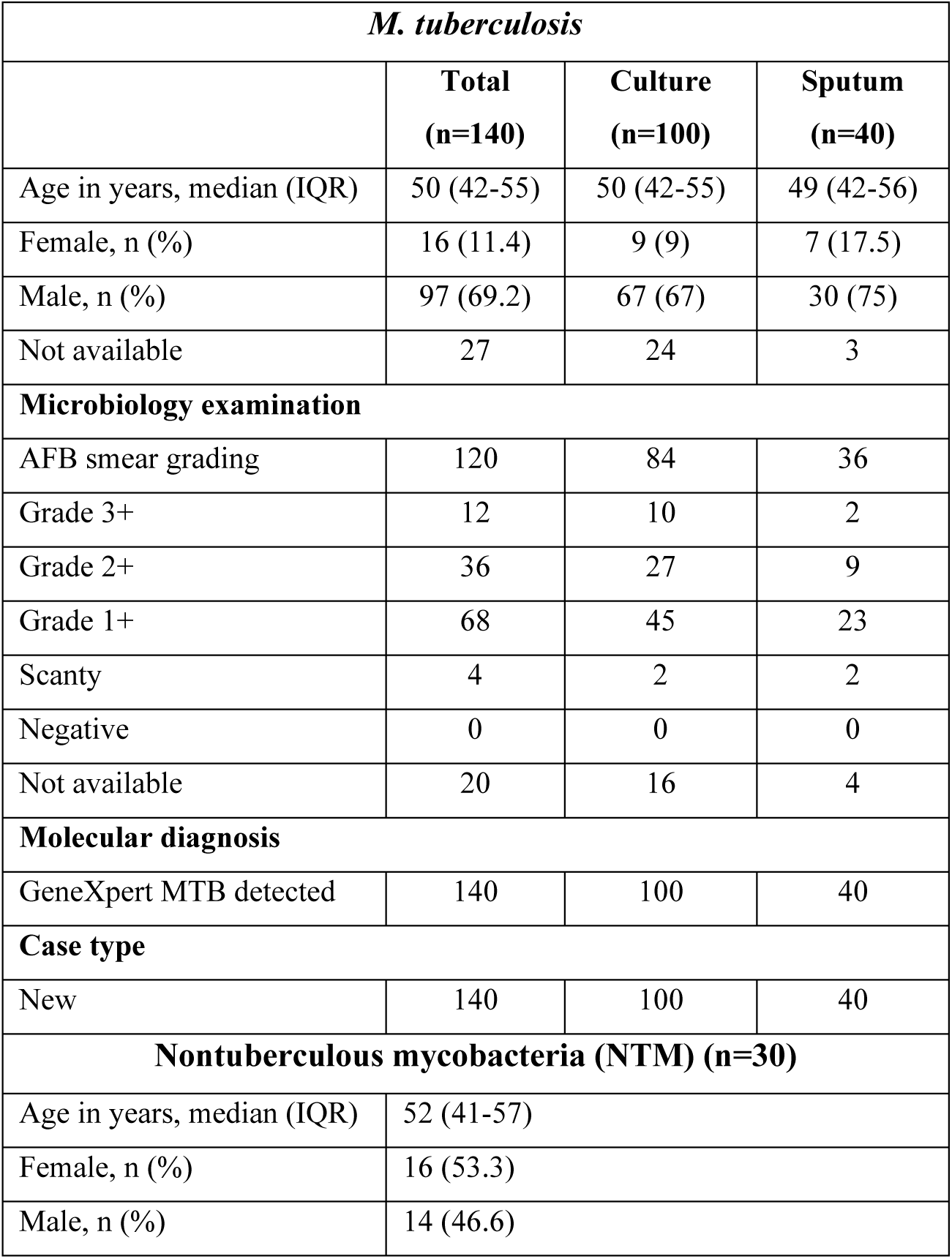
Clinical characteristics of the study cohort.

| *M. tuberculosis* |  |  |  |
| --- | --- | --- | --- |
|  | **Total**<br>**(n=140)** | **Culture**<br>**(n=100)** | **Sputum**<br>**(n=40)** |
| Age in years, median (IQR) | 50 (42-55) | 50 (42-55) | 49 (42-56) |
| Female, n (%) | 16 (11.4) | 9 (9) | 7 (17.5) |
| Male, n (%) | 97 (69.2) | 67 (67) | 30 (75) |
| Not available | 27 | 24 | 3 |
| **Microbiology examination** |  |  |  |
| AFB smear grading | 120 | 84 | 36 |
| Grade 3+ | 12 | 10 | 2 |
| Grade 2+ | 36 | 27 | 9 |
| Grade 1+ | 68 | 45 | 23 |
| Scanty | 4 | 2 | 2 |
| Negative | 0 | 0 | 0 |
| Not available | 20 | 16 | 4 |
| **Molecular diagnosis** |  |  |  |
| GeneXpert MTB detected | 140 | 100 | 40 |
| **Case type** |  |  |  |
| New | 140 | 100 | 40 |
| **Nontuberculous mycobacteria (NTM) (n=30)** |  |  |  |
| Age in years, median (IQR) | 52 (41-57) |  |  |
| Female, n (%) | 16 (53.3) |  |  |
| Male, n (%) | 14 (46.6) |  |  |

In addition, we included 30 NTM strains for assay validation. The median age of patients with NTM disease was 52 years (IQR; 41-57), and around 53% were women. Among NTM isolates, 75% (25/30) were identified as *Mycobacterium abscessus* and 25% (5/30) as *Mycobacterium fortuitum* (Supplementary table 1).

### Accuracy and analytical sensitivity of the probe

Amplification was assessed using *M. tuberculosis* H37Rv DNA as a positive control and a no template control. H37Rv replicates amplified with consistent Ct values, while the NTC remained negative, indicating high reproducibility and specificity. Melt-curve analysis showed a single peak (Tm=73.5°C). Serial 10-fold dilutions from 10^6^ to 10^1^ copies/reaction, tested in triplicate, confirmed reliable detection down to 10 copies/µL, establishing the limit of detection and demonstrating high analytical sensitivity (Fig. 2).

**Figure 2:**
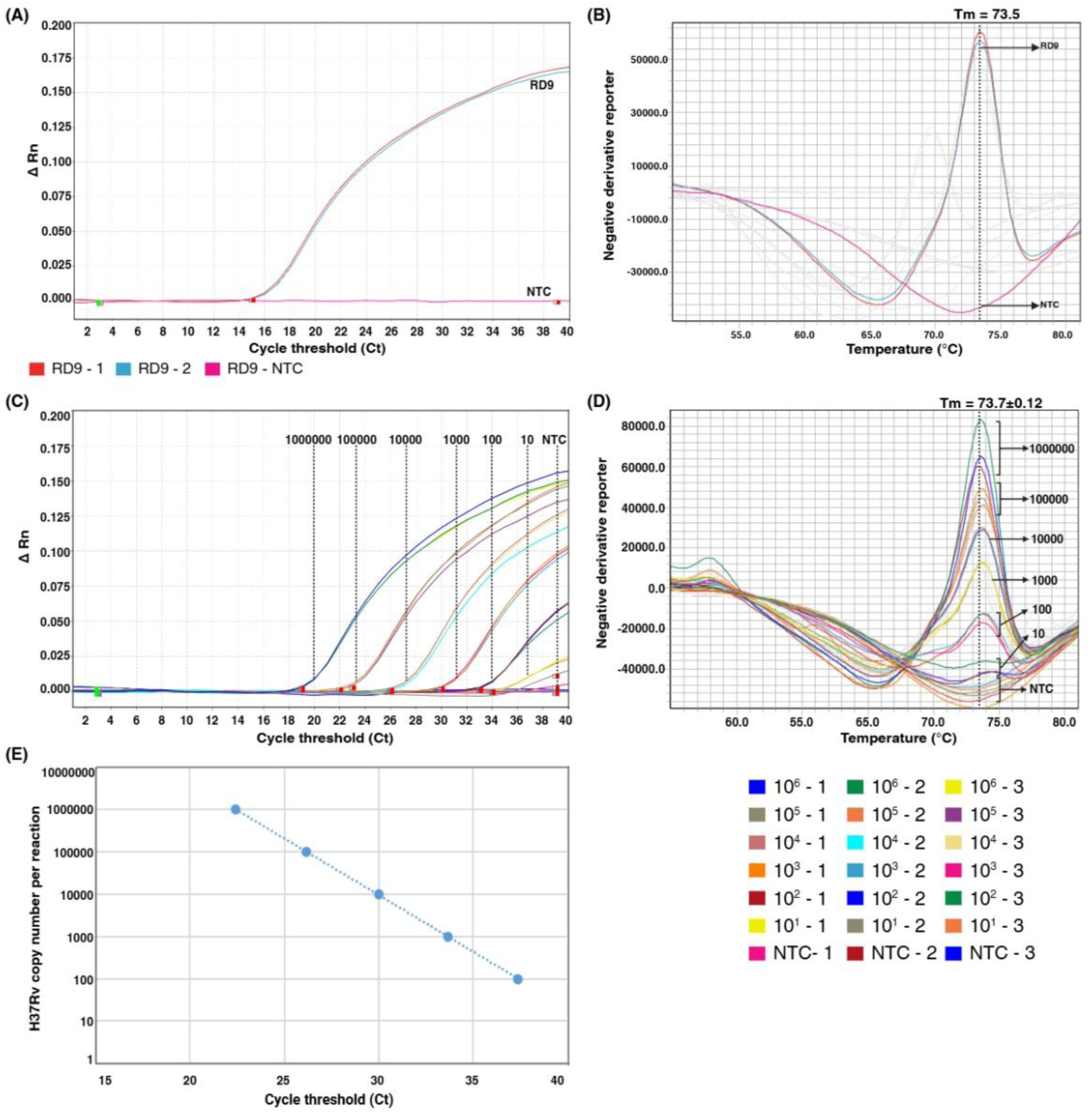
Accuracy assessment using *M. tuberculosis* H37Rv DNA and determination of the limit of detection. **(A)** Amplification plot of H37Rv replicates targeting the RD9 gene. **(B)** Corresponding melt curve showing a specific melting temperature (Tm) of ∼75°C. H37Rv DNA was serially diluted from 10^6^ to 10^1^ copies per reaction and analysed in triplicate. **(C)** Amplification plot obtained after 40 cycles for the dilutions, including the no-template control (NTC). **(D)** Corresponding melt curve showing a Tm of 73.7 ± 0.12°C. **(E)** Standard curve generated from the 10-fold serial dilutions, plotting cycle threshold (Ct) versus log copy number per reaction.

### Analysis to rule out non-specific binding

We performed the assay on saliva samples (n=10) from healthy controls with no clinical history of TB to confirm the absence of non-specific binding or false positive amplification of RD9 probe. Saliva was selected because it a complex biological matrix which contains diverse oral microbiota, human genomic DNA and PCR inhibitors making it a suitable matrix for evaluating potential non-specific binding. Out of 10 saliva samples tested, we did not detect any amplification and melt curve peaks up to 40 cycles. The absence of peaks indicated that no off-target amplification was observed for the developed assay. These results confirm that our probe is highly specific and does not react with non-target DNA in a complex biological matrix (Fig. 3A).

**Figure 3:**
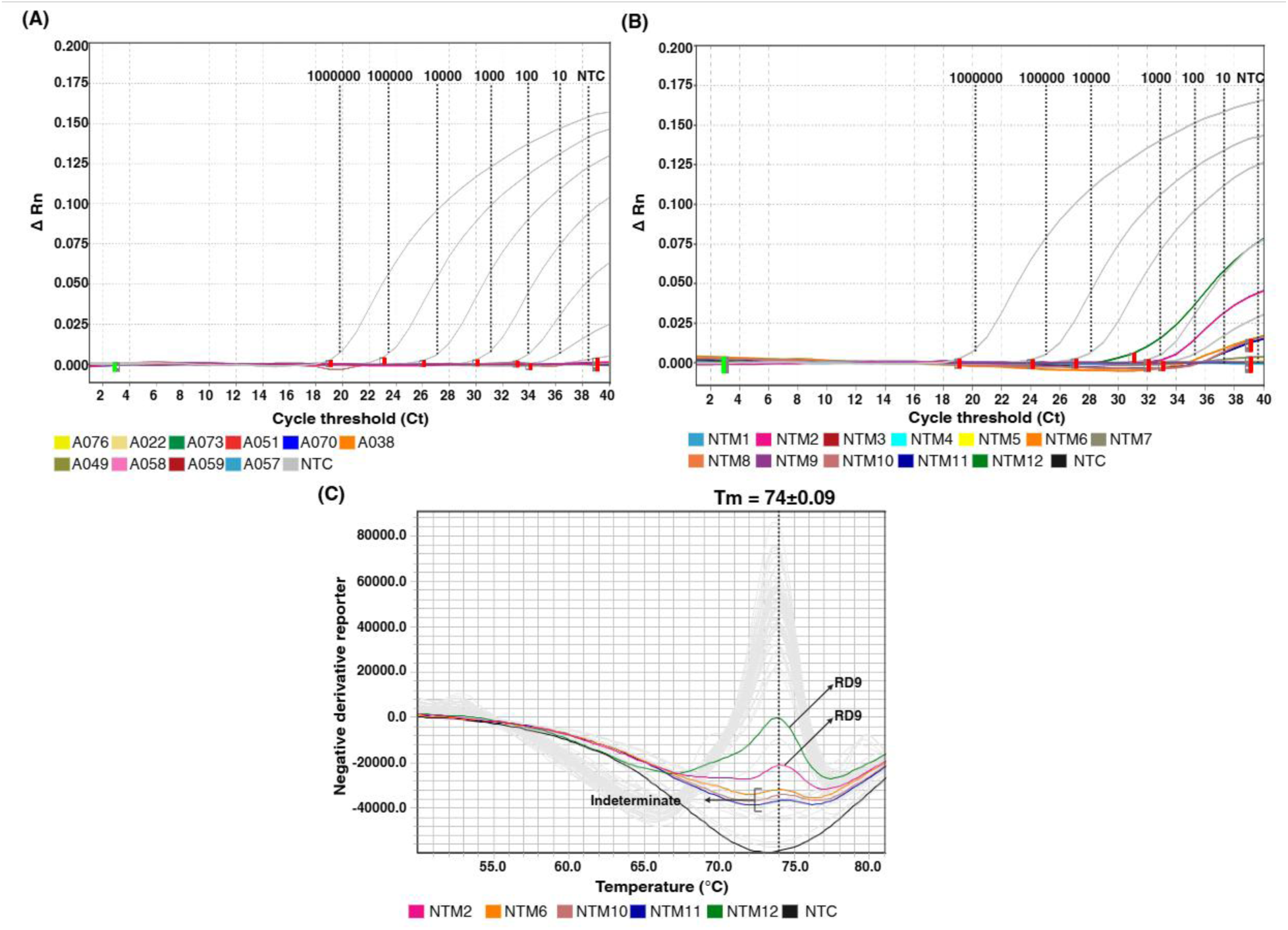
Specificity assessment of the RD9 probe for non-specific binding. Genomic DNA from 10 saliva samples of healthy controls was tested with the RD9 probe. **(A)** Amplification plot showing no amplification in all the samples. Additionally, 30 non-tuberculosis mycobacterium (NTM) isolates (*Mycobacterium abscessus and Mycobacterium fortuitum*) were analysed using the RD9 probe. **(B)** A representative amplification plot of NTM samples. **(C)** Melt curve plot of NTM samples showing Tm of 74 ± 0.09°C. NTC- No template control

### Probe specificity: *M. tuberculosis* Vs NTM

The validation cohort included 140 *M. tuberculosis* samples (40 sputum, 100 culture isolates) and 30 NTM culture isolates. Because some NTM share partial genomic similarity with *M. tuberculosis*, we specifically assessed RD9 probe reactivity against NTM. The NTM panel comprised *Mycobacterium abscessus* and *Mycobacterium fortuitum* isolates, neither of which contains the RD9 locus. In NTM samples, we did not observe any non-specific melt peaks, indicating high probe sensitivity. Amplification was detected in 6/30 samples, of which four had late Ct values of 39. Melt-curve analysis for these four samples showed a broad peak; therefore, they were categorized as “indeterminate.” Two samples showed amplification with a well-defined RD9-specific melt peak. Both samples were *Mycobacterium abscessus* culture isolates with *M. tuberculosis* copy numbers of 1141 and 216. These findings suggest possible *M. tuberculosis* and NTM co-infection, which has been reported previously by several studies. However, they could also reflect *M. tuberculosis* contamination during sample processing at the clinical site, as the NTC in all qPCR reactions was negative and the majority of NTM samples did not show any amplification (Fig. 3B and C).

### Clinical validation on culture isolates

For clinical validation, DNA was extracted from 100 *M. tuberculosis* culture isolates that were positive by both Xpert MTB/RIF and MGIT culture. The assay showed 100% concordance with culture and Xpert for *M. tuberculosis* detection, with a median Ct of 20.6 (IQR 19.4-22.3). Amplification was obtained in all culture-derived DNA samples (100/100). HRM analysis demonstrated a DNA-probe hybrid melt peak at 73.7±0.2°C in culture isolates, whereas standards analysed in the same run (10^6^ to 10 copies) showed a peak at 73.8±0.25°C, confirming accurate RD9 detection across samples. The assay LOD was 10 copies per reaction, at which *M. tuberculosis* genome copies were quantifiable in all culture isolates, with a median of 2,864,452 copies/reaction (IQR; 1,004,022-5,701,038) (Fig. 4).

**Figure 4:**
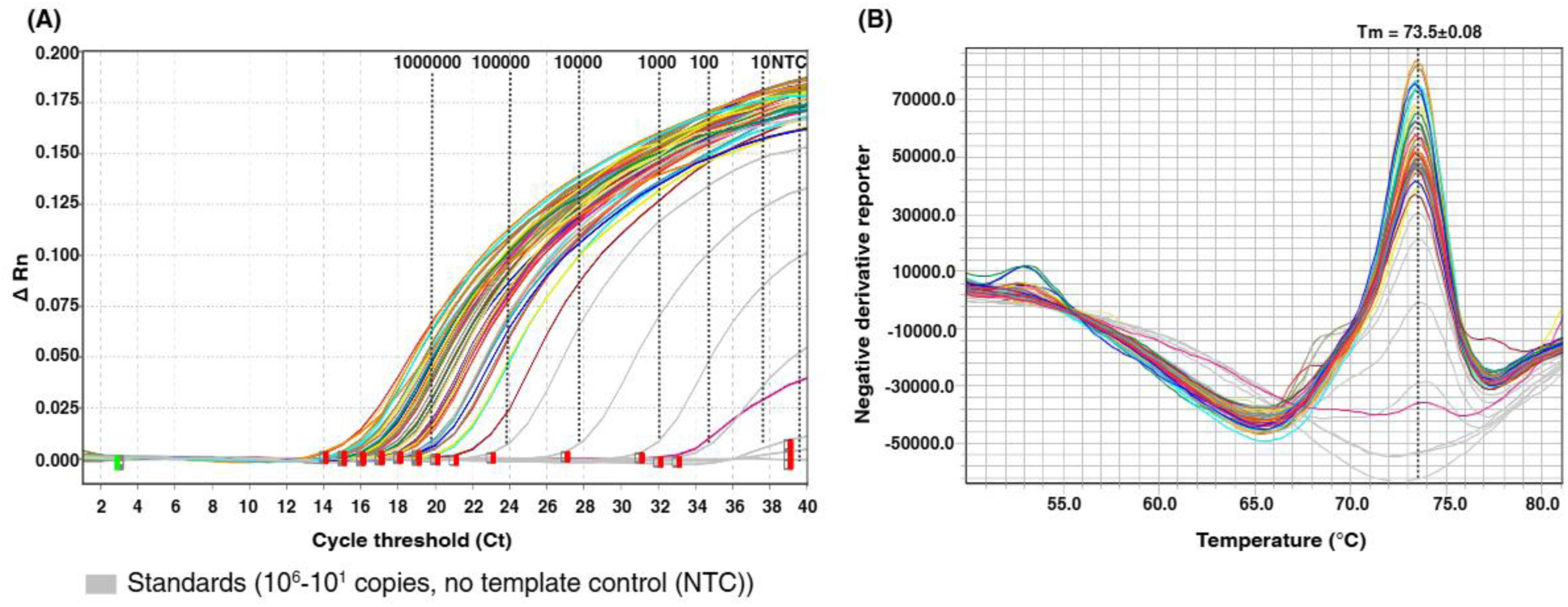
A representative figure for the clinical validation of the RD9 probe using 50 culture *M. tuberculosis* samples (out of 100 samples tested). (A) Amplification plot showing that all the culture samples amplified, and **(B)** the melting temperature (Tm) for these samples was 73.5 ± 0.08°C. Amplification and the melting curves of the culture isolates are represented with different colours, whereas the standards (10^6^-10 copies) and no template control (NTC) are shown in grey.

### Clinical validation on sputum samples

Sputum samples (n=40) that were Xpert MTB/RIF-positive were analysed using the HRM-qPCR assay. Amplification was obtained in 38/40 samples (95%), with a median Ct of 29.5 (IQR; 27.7-32.5). HRM profiles showed a consistent DNA-probe hybrid melt peak at 73.7±0.19°C in sputum DNA, closely matching the peak for quantification standards (74.0±0.08°C), which supports accurate RD9 target detection. At the assay LOD (10 copies/reaction), *M. tuberculosis* loads in sputum had a median of 3,367 genome copies per reaction (IQR; 576-11,971). To explore the relationship between bacillary burden and smear status, we analysed 36 samples with valid AFB smear grades and Ct values. Most samples were smear 1+ (23/36), followed by 2+ (9/36), with two Scanty and two 3+. Median Ct values by smear grade were 29 for 1+ and 2+ each, and 30 for 3+. Spearman’s correlation between smear grade and Ct was 0.29 (p=0.089), indicating a weak, non-significant inverse association (Fig. 5).

**Figure 5:**
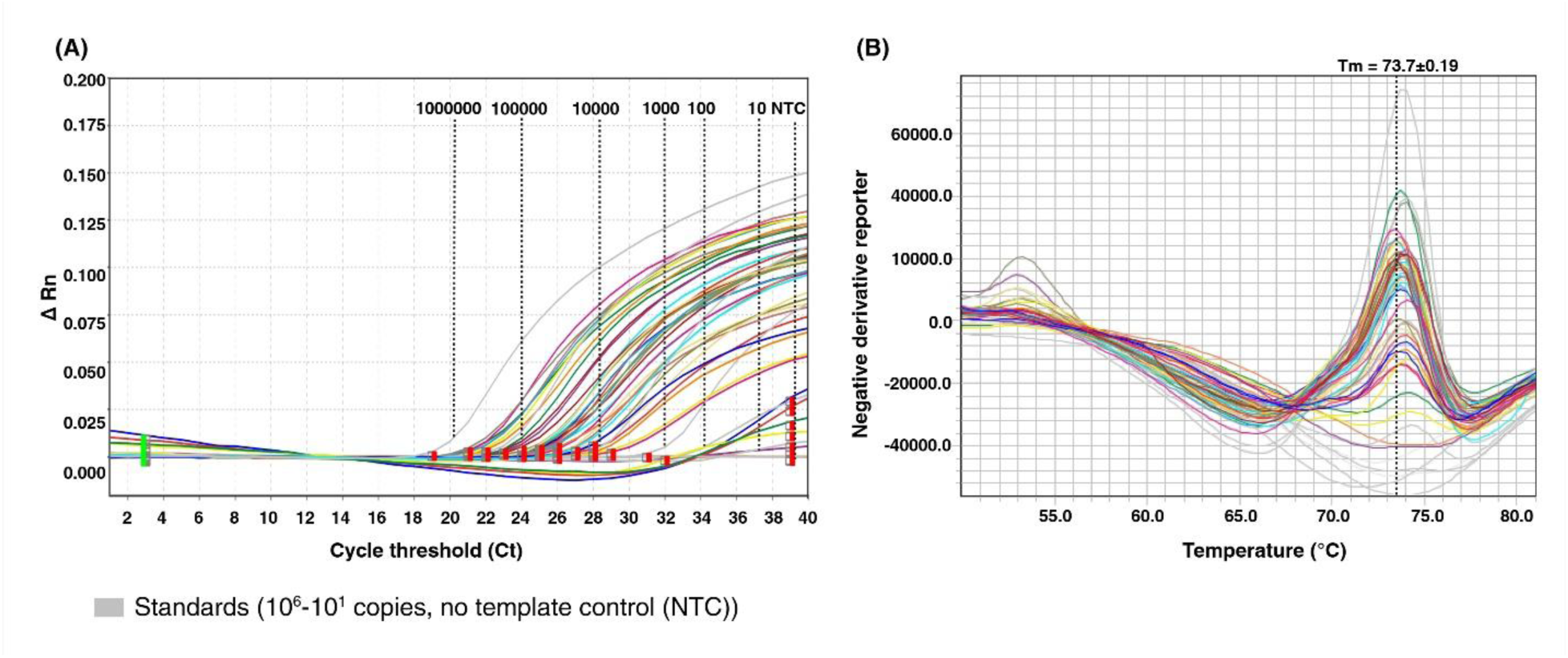
Clinical validation of the RD9 probe using DNA extracted directly from sputum samples (n=40). **(A)** Amplification plot showing successful amplification in 38 out of 40 samples (95%). **(B)** Corresponding melt curve analysis showing a melting temperature (Tm) of 73.7 ± 0.19°C across the positive samples. Amplification and the melting curves of the direct sputum samples are represented in different colours, whereas the standards (10^6^-10 copies) and no template control (NTC) are shown in grey.

## Discussion

Despite the recognized advantages of molecular bacterial load assays, rapid, affordable and scalable tools for quantifying bacillary load in routine clinical settings remain limited. Most assays currently used in TB diagnostics, such as Xpert MTB/RIF and TrueNat, are qualitative or at best semiquantitative, while commercially available quantitative assays are often restricted to research use because of their high cost. To address these gaps, we developed a rapid quantitative real-time PCR assay using molecular beacon chemistry and conducted comprehensive analytical and clinical validation. Our assay demonstrated 100% specificity and high sensitivity for detecting *M. tuberculosis* in both culture isolates and sputum samples, with no non-specific amplification observed in non-tuberculous mycobacteria. This assay reliably detected as few as 10 genome copies, and results were obtained in 90 minutes, underscoring its potential as a rapid and scalable tool for bacterial load estimation in high-burden settings.

Most quantitative *M. tuberculosis* assays currently in use are based on TaqMan chemistry [11,16]. While TaqMan probes are highly sensitive, several studies have shown that assay performance becomes more variable and error-prone at low copy numbers because of stochastic effects near the limit of detection. Low-titre clinical samples and sporadic late amplification for certain bacterial targets, therefore, warrant confirmatory testing and further prospective validation before broader implementation [17,18]. Incorporating an additional HRM step that generates a target-specific melt signature can help mitigate this limitation. In our assay, even at low copy numbers (n=10), late amplification events were accompanied by a characteristic melt profile, providing confirmation that the amplification signal is real, in contrast to conventional TaqMan assays where interpretation typically relies only on amplification curves and Ct values.

Several quantitative PCR assays for *M. tuberculosis* have targeted the insertion sequence IS6110, exploiting its multicopy nature to enhance analytical sensitivity and to infer bacillary load from Ct values [19]. IS6110 copy number, however, varies widely between strains, ranging from zero to more than 20 copies per genome and differing across lineages and even within the same lineage [20,21]. Comparative evaluations of single-copy versus multicopy qPCR targets have shown that although IS6110-based assays perform well for detection, they are less reliable for absolute quantification across genetically diverse strains and can under- or over-estimate bacterial burden when a fixed copy number per genome is assumed [19, 22]. In contrast, our assay targets the single-copy RD9 locus, for which one amplicon corresponds to one genome equivalent of *M. tuberculosis* [23]. By using this single-copy, *M. tuberculosis*-specific target together with molecular beacon chemistry and HRM confirmation, our assay provides lineage-independent absolute quantification and overcomes the key limitations inherent to IS6110-based quantitative approaches.

*M. tuberculosis*-NTM co-infection is relatively uncommon but clinically important, with regional series reporting NTM in 2-8% of patients diagnosed with TB or TB suspects [24–26]. A multicentre study from China reported *M. tuberculosis*-NTM co-infection in approximately 3% of patients evaluated for MDR-TB. Since NTM isolates are frequently resistant to first-line anti-TB drugs, such mixed infections may be misdiagnosed as MDR-TB in high-burden countries [27]. Most routine molecular assays are optimised for either MTBC or NTM and are not designed to detect low-level *M. tuberculosis* in specimens dominated by NTM. In our study, the RD9-targeted molecular-beacon qPCR could reliably detect *M. tuberculosis* DNA against a high NTM background as each *M. tuberculosis* signal is independently confirmed by an *M. tuberculosis*-specific HRM profile, reducing the risk of false-positive late amplification. Commercial multiplex assays such as the Anyplex MTB/NTM real-time detection test can simultaneously detect and differentiate MTBC and NTM in respiratory samples, with *M. tuberculosis* sensitivities of about 80-90% in smear-positive disease, but they largely provide qualitative results and do not systematically characterise minor *M. tuberculosis* subpopulations in mixed infections [28]. Multiplex PCRs targeting genes such as *rpoB* or *hsp65* show similar constraints, often requiring sequencing to confirm true co-infection [29]. By using a single-copy, *M. tuberculosis*-specific RD9 locus combined with HRM confirmation, our assay offers higher analytical sensitivity for minor *M. tuberculosis* subpopulations and built-in verification of co-infection, positioning it as a complementary tool to existing *M. tuberculosis*/NTM platforms. Another major application of our RD9 HRM assay is to define the minimum bacillary load required for targeted sequencing. In the multicountry Seq&Treat tNGS implementation study, valid sequencing results were obtained for 88.5% (GenoScreen) and 93.1% (Oxford Nanopore Technologies) of 763 DR-TB samples; however, sequencing succeeded in only 2-8% of specimens in the “very low” Xpert category compared with 29-43% in the “low” category, indicating frequent failure in paucibacillary samples due to low *M. tuberculosis* DNA or poor library performance [30]. A culture-free tNGS evaluation similarly reported good performance only for specimens with moderate or high bacterial loads, while many low-load samples failed or yielded incomplete resistance profiles, underscoring limited utility in paucibacillary disease [31]. Integrating our quantitative HRM-qPCR into existing tNGS workflows to pre-screen specimens for a minimum *M. tuberculosis* genome copy number could reduce failed sequencing runs and optimise use of sequencing resources.

Our study has several limitations. We evaluated the HRM assay in a limited number of sputum samples, all of which were AFB-positive. Larger cohorts that include paucibacillary sputum specimens will be required to confirm the limit of detection in clinical samples. Although our assay can detect *M. tuberculosis* and NTM co-infection, we did not perform any spiking experiments to evaluate the limit of detection of minority *M. tuberculosis* co-infection. Additionally, we tested only 2 NTM strains (*Mycobacterium abscessus* and *Mycobacterium fortuitum*) to assess the cross-reactivity of the probe. Finally, although we compared our assay with Xpert MTB/RIF and culture, we were unable to perform a full head-to-head evaluation against existing quantitative *M. tuberculosis* load assays due to funding constraints.

## Conclusions

In conclusion, the HRM-qPCR assay demonstrated a broad dynamic range, high analytical sensitivity with a 10-copy limit of detection, and excellent concordance with culture and Xpert MTB/RIF for the detection of *M. tuberculosis* DNA. HRM profiles were highly reproducible, with tight clustering of melt peaks between standards and clinical specimens, confirming robust target specificity. No amplification was observed in NTM or saliva controls, indicating high analytical specificity for *M. tuberculosis* even in complex biological matrices. Quantitative results from sputum samples showed that genome copy numbers are measurable across a clinically relevant range and broadly reflect smear grade-related differences in bacillary burden, although the correlation between smear grade and Ct was modest in this cohort. Collectively, these findings support the HRM-qPCR RD9 assay as a rapid, affordable tool for standardized *M. tuberculosis* load quantification, with potential applications in treatment monitoring, sample triage for sequencing, and transmission-risk assessment in high-burden settings.

## Supporting information

Babu et.al_Supplementary table 1

## Acknowledgements

The authors acknowledge the technical staff and patients from the clinical site for their cooperation during the study.

## Author Contributions

RV and NJM conceived and designed the study. VG and SVK enrolled patients and collected clinical data. KE performed DNA extraction, culture and LPA. RV and ASB designed and validated the HRM-qPCR assay. ASB performed the HRM-qPCR experiments. PY and PP analysed the data and prepared the tables and figures. RV and ASB drafted the initial manuscript. NMJ and AP contributed to data analysis and interpretation. All authors reviewed, revised, and approved the final manuscript.

## Data availability

The data that support the findings of this study are available on request from the corresponding author.

